# Heterogeneity in SIR epidemics modeling: superspreaders

**DOI:** 10.1101/2020.07.02.20145490

**Authors:** Istvan Szapudi

## Abstract

Deterministic epidemic models, such as the Susceptible-Infected-Recovered (SIR) model, are immensely useful even if they lack the nuance and complexity of social contacts at the heart of network science modeling. Here we present a simple modification of the SIR equations to include the heterogeneity of social connection networks. A typical power-law model of social interactions from network science reproduces the observation that individuals with a high number of contacts, “hubs” or “superspreaders”, can become the primary conduits for transmission. Conversely, once the tail of the distribution is saturated, herd immunity sets in at a smaller overall recovered fraction than in the analogous SIR model. The new dynamical equations suggest that cutting off the tail of the social connection distribution, i.e., stopping superspreaders, is an efficient non-pharmaceutical intervention to slow the spread of a pandemic, such as COVID-19.

## Introduction

According to recent results on the spread of COVID-19, a small fraction of the population is responsible for most infections (1, 2). Clusters in care facilities, restaurants and bars, workplaces, and music events (3), or even choir practice (4), dance events (5) are a hallmark of superspreading (6). The most wide-spread deterministic Kermack-McKendrick equations (7), or SIR equations, are not designed to model superspreading. Superspreading is introduced as an additional dispersion of the secondary infections (6) using a negative binomial distribution, possibly in the context of random network theory (8), or modeled with stochastic Markov Chain methods (9).

The simplest way to modify the SIR equations to include superspreaders is by adding a new class of susceptible individuals, P, superspreaders (10) to the equations. However, network science (11) suggests a more complex picture. If we map individuals into vertices of a graph and their connections into edges, their degree distribution, i.e., the number of social connections of an individual, typically follows a power-law distribution with a slope between 2 and 3. Recent analyses COVID-19 data (12) support such power-law behavior that is the hallmark of small-world phenomena of network science. Our goal is to use network science insight to generalize the SIR formalism. The result is a set of equations similar to poly-SIR generalizations of the SIR formalism (13, 14), but more specific to network science and does not assume a linear contact matrix: non-linearity is essential to superspreading. Note that here we are focusing on *structural* superspreading due to the average contact structure of the population as they go about their daily lives. In the discussions, we will outline how to model *transient* superspreading, i.e., rare individual events that can dominate the case counts when the overall numbers are comparable to the cut-off of the contact (degree) distribution, and *viral* superspreading, from those (if they exist) who shed more virus than average.

The principal idea is that we replace the susceptible-infected-recovered variables with a (binned) distribution in terms of their contacts. In the next section, we derive the equations governing the time development of these distributions; in section 3, we illustrate the dependence of the solutions of the network science parameters; and in the final section, we summarize and discuss the results.

### Network science inspired generalization of the SIR equations

Let us denote the number of social connections or contacts of a person with *k*. If a person is represented as a vertex of a graph, *k* would be the number of links of that vertex to other persons, or the degree of that vertex. Note that in reality this graph is changing every day: for instance, a person going to a rock concert would have many contacts on a particular day. We assume that over time, the contact distribution itself is stationary at least over the fundamental timescales associated with the disease. This is an approximation that will smear out transient rare events and can be fixed only with Monte Carlo modeling briefly discussed at the end.

The probability per day that a contact from an infected person produces a new infection is *β* = 1*/T*_*c*_, or the inverse of the average time in days that such connections will produce a new infection. In the usual SIR type modeling, the time derivative 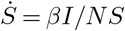, i.e., the infected fraction of the population *I/N* times the *S* chances of infections with probability *β* per day that an encounter results in a successful transmission. The original interpretation of *β* incorporates social interactions, while we split it off to do social contact distributions defined next.

To model the social connection network without a Monte Carlo representation of the actual social connection graph, we introduce the quantities, *S*_*k*_, *I*_*k*_, *R*_*k*_, the susceptible, infected, and recovered number of the population with *k* social links. The sum of the variables, *S*_*k*_ + *I*_*k*_ + *R*_*k*_ = *N*_*k*_ *∝k*^*−α*^ typically from network science. 2 *< α <* 3 (“ultra small world”) is the most interesting and practical limit for social networks. *α <* 2 is pathological, and *α >* 3 corresponds to random graph (“small world”). Typical ultra small world social networks contain large “hubs”, individuals with large number of connections. They are positioned in the tail of the distribution, and they will correspond to superspreaders in an epidemiological network.

With the above definitions we will write the network science inspired modification of the SIR equations as

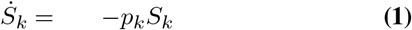

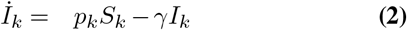

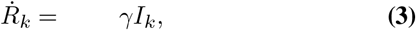

where *p*_*k*_ is the probability that a vertex with *k* links will be infected, implicitly depending on *I*_*k*_ and *β*, while *γ* = 1*/T*_*r*_ is the usual inverse recovery time in unit of 1/days. Note that the sum of any variable over index *k* gives the corresponding traditional SIR variable, and the sum of the equations correspond to an effective SIR equation with changing parameters. The above equations will describe how an initial distribution *S*_*k*_ responds to the disease dynamics. We estimate *p*_*k*_ next. It corresponds to the chance per day that a susceptible person with *k* social links will get infected. It is equal to 1 − *q*_*k*_, where *q*_*k*_ is the probability that the person in question does not get infected during that day, i.e. (1 *βp*)^*k*^, where *p* is the probability that one random link carries an infection (here we assume no correlation between infected links, a zeroth order approximation). Finally, in the spirit of the original SIR approximation, we approximate the probability *p* with the ratio of the infected links to the total number of links. Putting all these together

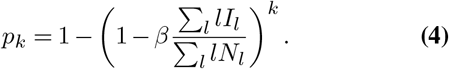

Note that we double counted both the infectious and total number of links which cancels in the ratio. While this is the simplest approximation for *p*_*k*_ and it does not account for topological details and degree correlations of the graph representing social interactions. The above random, non-associative network assumption captures many features of network theory without a full graph Monte Carlo simulation. The sum of these variables over *k* will obey an effective SIR equation with

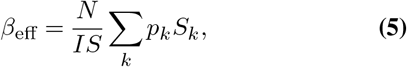

with *N* = ∑_*k*_ *N*_*k*_, etc. Initially, the effective *R*_0_ = *β*_eff_ */γ*, i.e., it depends on the social dynamics through the underlying *k* dependence and the disease dynamics represented by *β*, and *γ*. Therefore a given pandemic, like COVID-19, could play out differently depending on the social interaction hierarchy of people.

## Results

We implemented the above model as a `python` code. If only the *k* = 1 terms are different from zero, then *p*_*k*_ = *βI/N*, and *β*_eff_ = *β*, i.e. our model is identical to the original SIR model. As a sanity check, we verified that this is the case.

To illustrate the flexibility and the qualitative behavior of the new model, we choose a set of fiducial (reference) parameters and plot solutions with respect to changing each parameter. First, we select a traditional SIR model as a baseline: we choose *β* = 0.2 and *γ* = 0.1, which corresponds to *R*_0_ = 2. The typical behavior of this SIR model is shown on Figure 1 with dotted lines.

**Fig 1.**
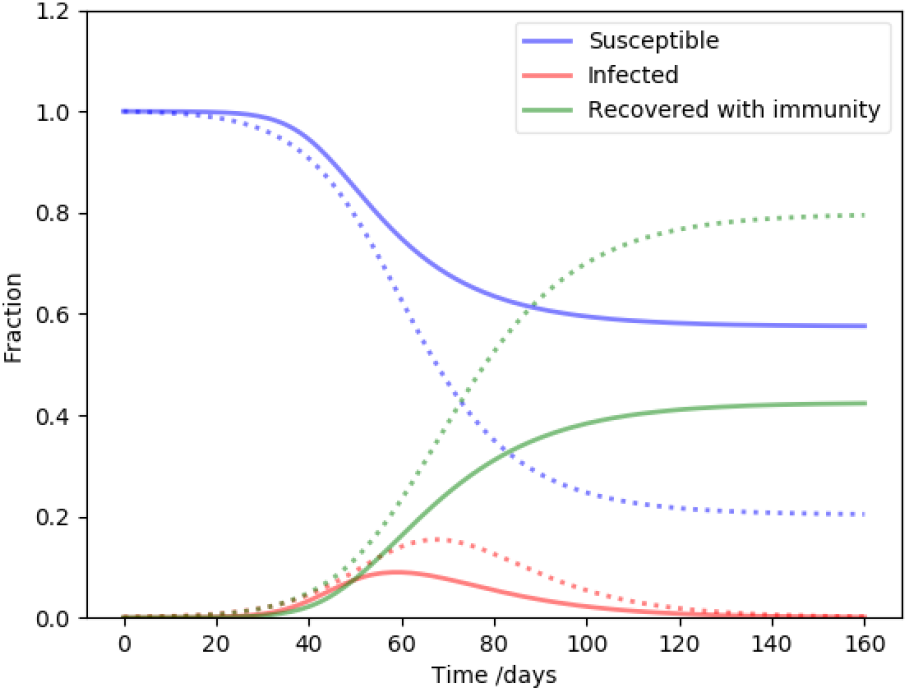
The dots present a typical traditional SIR model run with *β* = 0.2, *γ* = 0.1 (*R*_0_ = 2), and one initial infection. The solid lines correspond to *β*_eff,0_ ≃ 0.7, i.e *R*_0_ ≃ 7, *k* = 1…30, one infection for a superspreader in the *k* = 10 bin, and *α* = 2.5 for the initial distribution. This model is our fiducial NSIR model and it is displayed with dots for reference the set of epi curves that follow.

For the network inspired model, labeled NSIR, we choose *β* = 0.07, *γ* = 0.1, a slope of the vertex degree distribution *α* = 2.5, the maximum number of connections (i.e. the cut-off of the vertex degree distribution, the largest super-spreader) *k*_max_ = 30. In this case initially *β*_eff_ ≃ 0.7, or *R*_0_ ≃ 7; this is much higher than the corresponding *R*_0_ = 2 in our baseline SIR model. As illustrated in Figure 1, even with this seemingly extreme initial condition, the time development of the disease dynamics is a milder version of the baseline SIR model. The peak infection rate is smaller, and a similarly smaller recovered fraction leads to herd immunity. This is a generic feature of models with significant super-spreaders: since they are responsible for a large fraction of infections, but also prone to get infected quickly, they eliminate themselves from the susceptible pool early on. Once that happened, the transmission is no longer efficient. As we will show later, (Figure 5), this process expresses itself as a precipitous drop in *R*_*t*_.

Next we demonstrate the behavior of the solutions in terms of individual parameters. We focus on the parameters describing the social network properties, the novel feature of our model. First, we take a look at the effect of inserting the infection at a particular degree node. Figure 2 demonstrates, the effect of introducing the infection at the highest or lowest degree node is relatively minor: infecting a highest degree individual (or hub) slightly speeds up the time-line, while infecting a low degree node slightly slows it down. Aside from a slight displacement of the peak, the effect is insignificant. Most importantly, the herd immunity level is not influenced by the transient behavior due to the insertion degree, therefore the conclusions that follow should be robust against that. Next we examine the cut-off of the degree distribution, corresponding to the highest degree hubs or superspreaders present in the population. While in the fiducial model we assumed at most *k*_max_ = 30 connections, Figure 3 shows 11 (top) and 100 (bottom) connections. The higher the cut-off, the faster the time development and the higher the peak infected rate is. Conversely, cutting off the superspreaders flattens the curve, as expected. The number of equations is three times the highest degree node, for *k*_max_ = 11, 30, 100, respectively, 33, 90, 300 coupled differential equations were solved.

**Fig 2.**
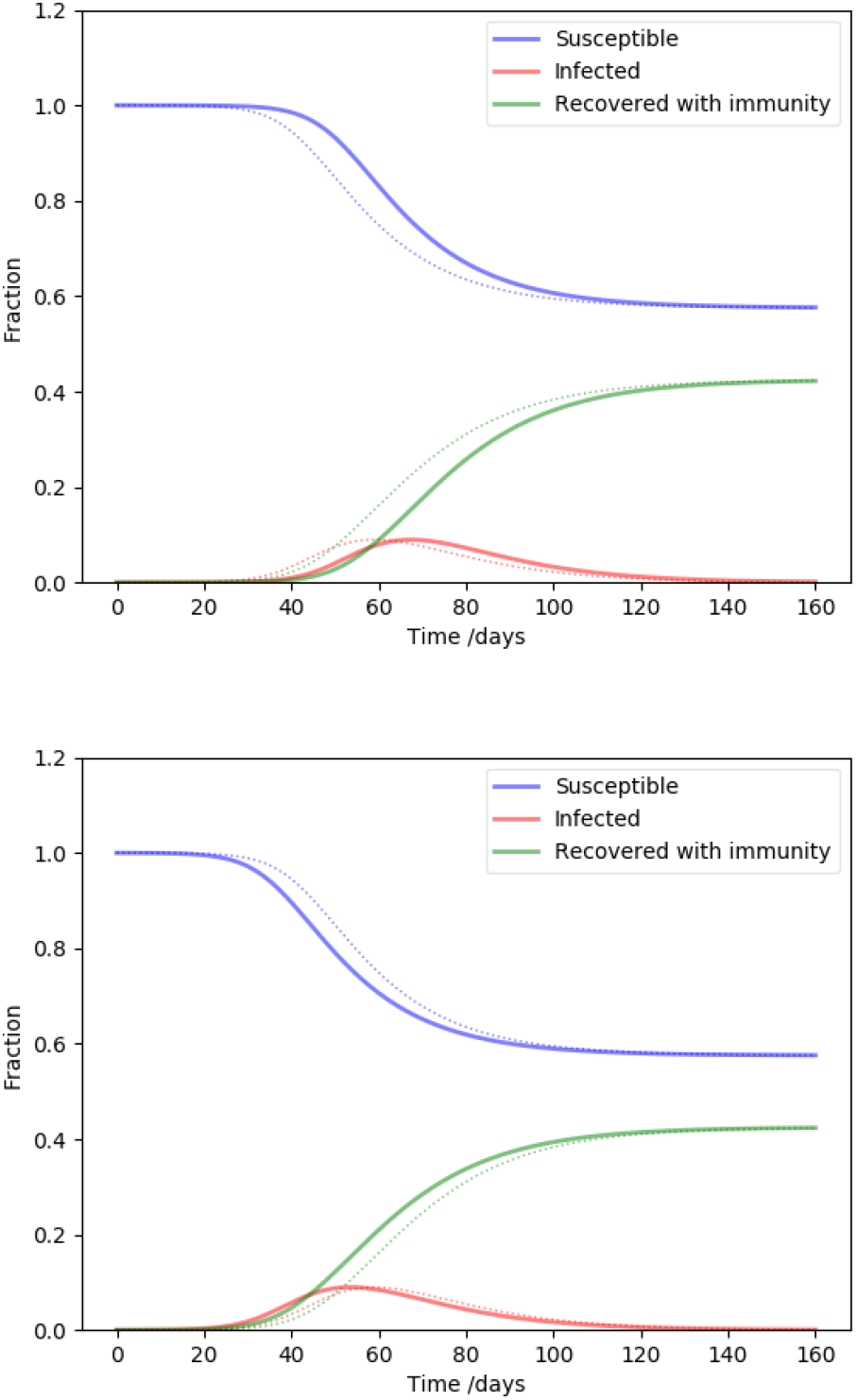
The effect of infecting nodes with different degrees. The parameters are the same as NSIR model of Figure 1, except the infection is inserted at the highest degree (top), and the lowest degree (bottom) node. The fiducial model is plotted with dots for reference.

**Fig 3.**
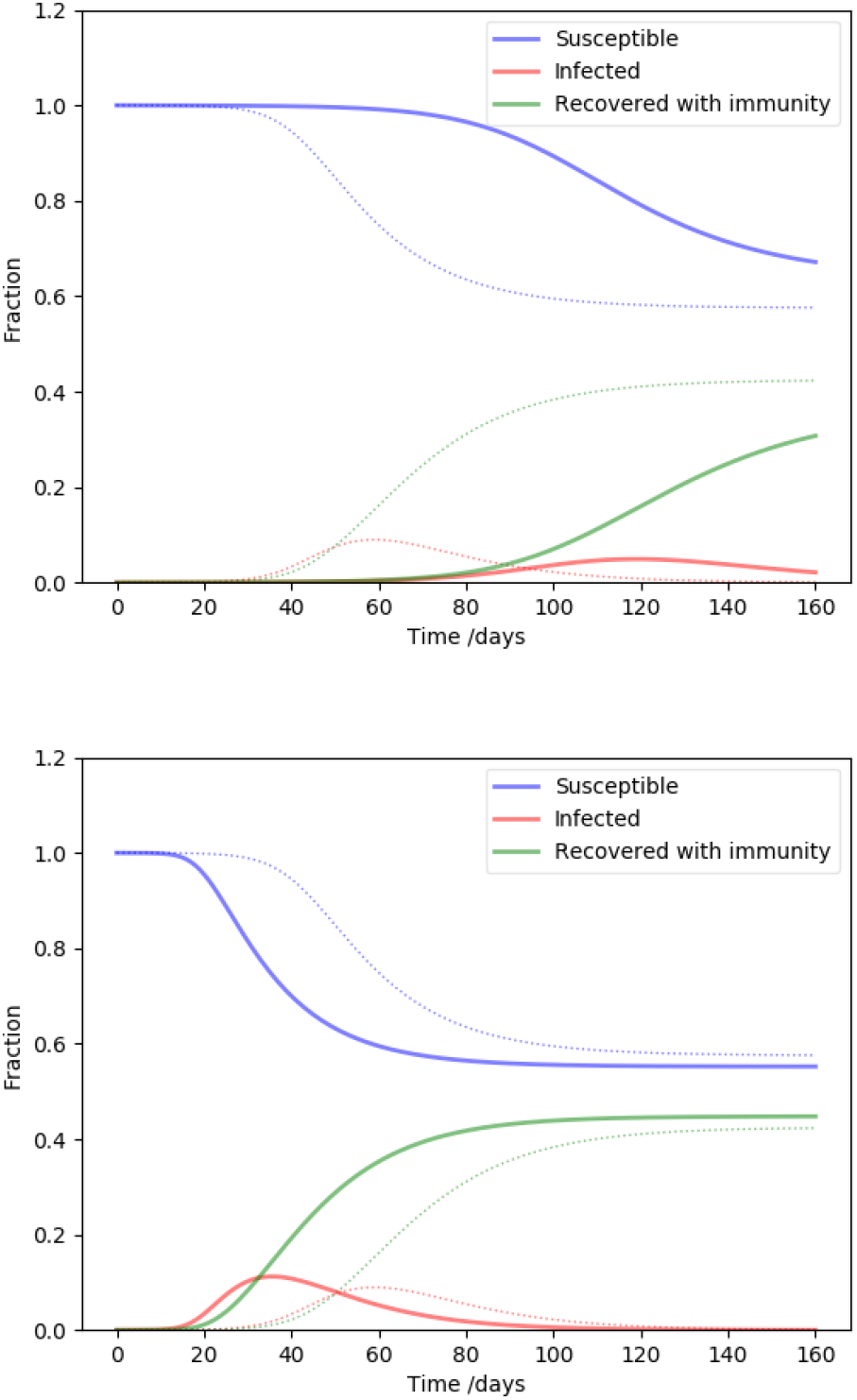
The effect of the highest degree nodes (the cut-off of the degree distribution). The parameters are the same as NSIR model of Figure 1, except the highest degree is 11 (top), and 100 (bottom). These two cases represent extreme, or moderate superspreaders, respectively. The fiducial model is plotted with dots for reference.

**Fig 4.**
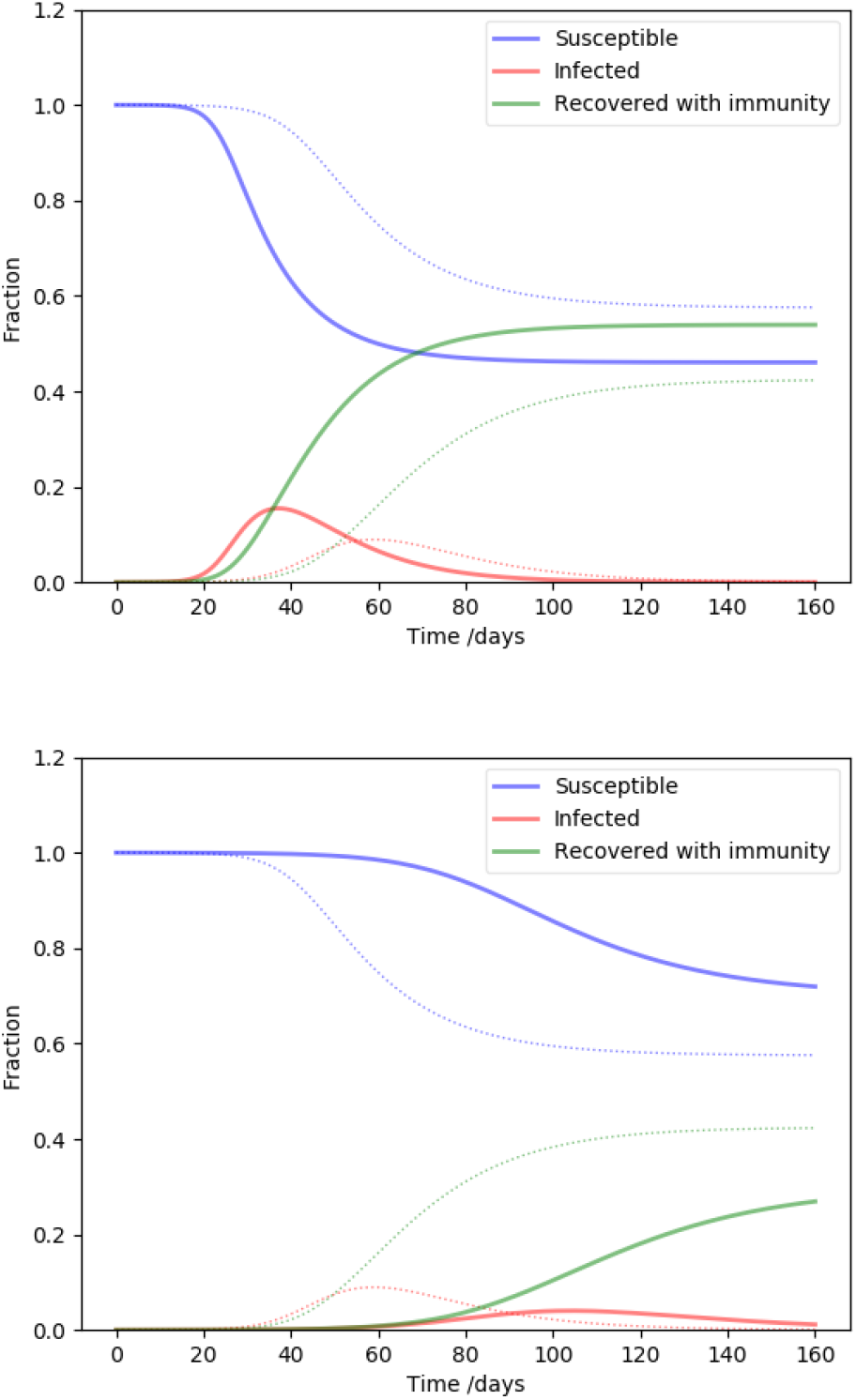
The effect of of the slope of the degree distribution. The parameters are the same as NSIR model of Figure 1, except *α* = 2.1 (top), and *α* = 2.9 (bottom). The flatter distribution means more superspreaders/hubs, while the steeper distribution corresponds to a case more similar to a random graph. The fiducial model is plotted with dots for reference.

**Fig 5.**
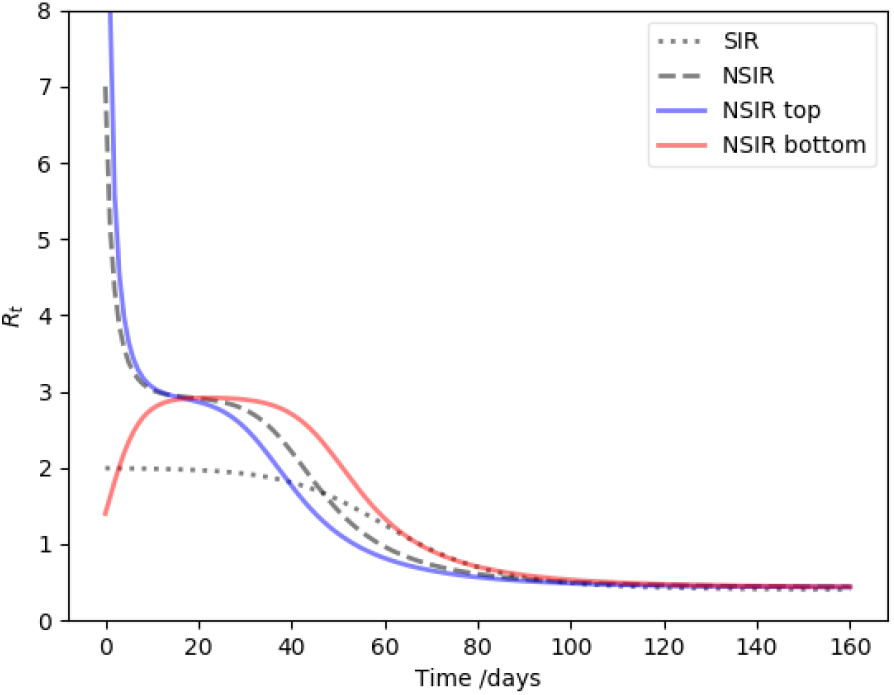
The time development of *R*^*t*^ is plotted for the fiducial SIR (dots), fiducial NSIR (dashes), and two extreme NSIR models where the infection is inserted into a high degree node (top, blue), or a low degree node (bottom, red). Independent of the insertion degree, each NSIR curve lingers around *R* ≃ 3. This is more meaningful than the actual *R*_0_ that is sensitive to the insertion degree.

The slope of the degree distribution controls the relative contribution of the rare high degree nodes (hubs, superspreaders) compared to the more usual low degree nodes. Our fiducial parameter *α* = 2.5 is middle of the range, and Figure 3 values close to the extremes in the interesting range from network science: *α* = 2.1 (top), corresponding to a distribution with a heavy tail, or many superspreaders, while *α* = 2.9 (bottom) is approaching a random network (*α* ≥ 3). It is clear that increasing the proportion of superspreaders increases the severity of the disease time-line, as before.

As we have shown earlier, whether the infection is introduced at a high degree or low degree nodes corresponds to transient effects, and hardly influences the top line final results. To elucidate the difference it makes in the early stage of the endemic, Figure 5 plots *R*_*t*_ for several cases: the fiducial SIR model, our fiducial NSIR model, and two extremes, where the infection was introduced at a hub (superspreader) node, or a bottom (low social connection) node. Eventually, all three NSIR models converge to the same SIR model asymptote. If the infection starts with a hub, the early values of *R*_*t*_, corresponding to an effective *R*_0_, are extremely high and drop quickly when the superspreaders eliminate themselves from the susceptible pool. Conversely, if the infection starts at a low degree node, the initial *R*_0_ is much lower, even rises initially as the infection spreads towards superspreaders. Therefore, if this is a realistic model of disease transmission, measurements of *R*_0_ are fairly sensitive to serendipitous transient effects due to where exactly the infection was introduced.

Finally, we show time development of each bin in the contact distribution of the population in our fiducial NSIR model. On Figure 6, the solid red lines from top to bottom correspond to the degree distribution at a particular point in time from low to high degree. For reference, the total SIR variables are displayed with dots on this semi-log plot: the red dots correspond to the sum of the red solid lines.

**Fig 6.**
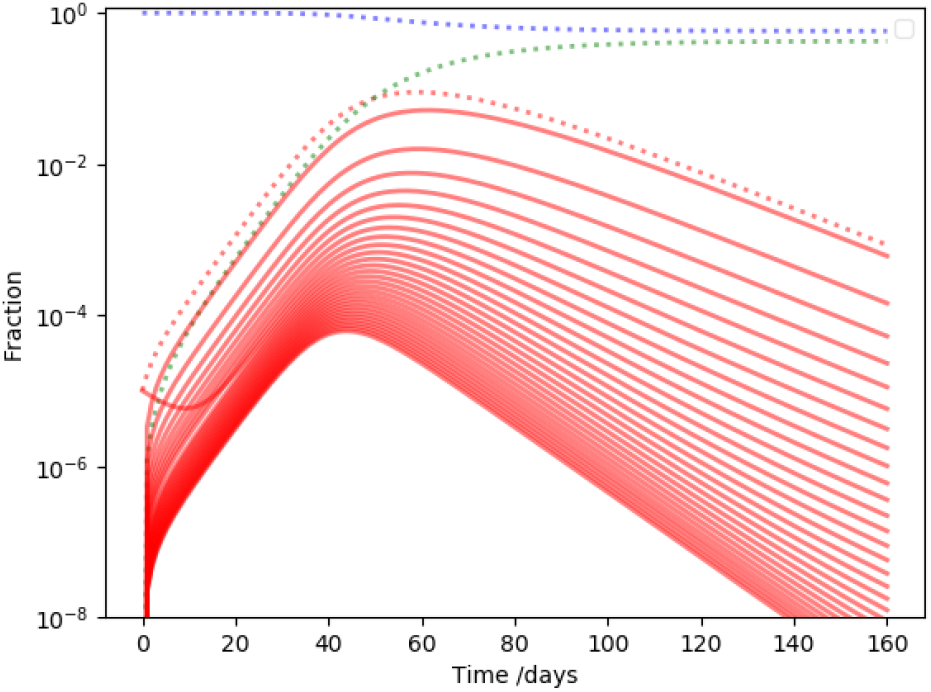
The time development if individual infection bins in the fiducial NSIR model. The dotted lines correspond to the summary of the fiducial model, while the red solid lines correspond to the individual bins if the infected distribution, the highest degree being the lowest point. The one red line with different transient behavior than the rest corresponds to the node where the infection was inserted.

After transient effects, nodes with different degrees have similar development, perhaps high degree nodes, hubs or super-spreaders, reaching their maxima slightly earlier. The one outlying solid line corresponds to the transient behavior of the node carrying the initial infection. After the first few weeks (while the infected fraction is still fairly low), the transient behavior joins the trend carved out by the rest of the curves. It took about two timescales of 1*/γ* to achieve such statistical “thermalization” and erase the memory of where the infection started.

## Summary and discussions

We have introduced a generalization of the SIR model, where each variable is a distribution. This generalization is a specific case of a poly-SIR model that could describe a variety of situations, e.g., (13, 14) used it to model the interactions of different age groups with a constant contact matrix. We focus on social connection distributions motivated by network science. The resulting equations are more non-linear than a standard poly-SIR model with a fixed contact matrix.

There are a number of phenomenological parameters that influence the dynamics of the disease transmission. In our formulation, both parameters *β* and *γ* are describe average properties of the viral infection, unlike the traditional SIR model, where *β* is influenced by the properties of the disease as well as social interactions. We model social interactions through the social contact distribution motivated from network science. The universality of social networks (11) motivates the assumptions of a power law degree distribution. We explore how the slope, the cut-off, and the contact degree of the initial infection drives the dynamics.

Our main result, consistent with expectations and results from other methods, is that if supserspreaders, highly connected hub-nodes, are important for disease transmission, a lower level of infection rate leads to herd immunity than otherwise. Superspreaders tend to infect each other quickly and remove themselves from the susceptible pool earlier. Thus, the initial effective *R*_0_ drops quicker than in the analogous SIR model.

The steeper the degree distribution and the lower its cut-off, the less important superspreaders become. Steepening the distribution, i.e. moving people to lower connections than normal, corresponds to social distancing or lock-down. Moving the cut-off to lower degrees, i.e., removing superspreaders appears to be an effective strategy to flatten the curve as well. If parameters are fit from realistic data, our results could inform non-pharmaceutical mitigation strategies during the COVID-19 pandemic.

We found that the insertion of the infection at a highly or poorly connected node results widely varying effective *R*_0_ values if interpreted in terms of the traditional SIR model. We speculate that zoonotic viral transmission to humans is likely at an average node (because there is more such nodes), while transmission by or after travel is more likely through hub nodes. This is qualitatively consistent with the initial lower estimates of *R*_0_ ≃ 2 *−* 3 for COVID-19 (15) that was revised up to *R*_0_ ≃ 4 *−* 6 (16, 17) after international spread.

Our work does not take into account the discrete transients of disease transmission: a particular person (or the corresponding node) is either infected or not, there are no fractional infections. In the contrary, the solutions to SIR-like differential equations have a scaling property: a new solution is obtained by multiplying a solution with an arbitrary real number. This is clearly not a good approximation for smaller communities and stochastic effects are important in the early phases of an endemic in larger communities. For instance, the first infection might die out or trigger a transient superspreader event, ultimately leading to vastly different outcomes.

It would be relatively straightforward to modify our model to include transients in a Monte Carlo fashion. Instead of ultiplying with the probability *p*_*k*_, we could draw the number of infections from a multinomial distribution of *S*_*k*_ total numbers with *p*_*k*_ infection probability, assuming each infection is independent. Our differential equations then become a discrete difference equations: 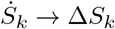, and so forth. Each solution corresponds to a Monte Carlo realization, and rare events, corresponding to mass gatherings, for instance, could now happen in a discrete fashion. If it is found that disease transmission probability varies widely with individuals *β* itself could be a realization from a distribution. If that distribution has a long tail, this process would induce viral super-spreading.

Several of the Monte Carlo simulations are needed to evaluate the statistics and small number transients associated with the transmission. Nevertheless, such Monte Carlo simulations would still be less costly than a full network science Monte Carlo model. Once the total numbers are high enough, the multinomial distribution will narrow enough to produce results very similar to our equations 3. The above generalizations are left for future work.

## Data Availability

All data in this article has been generated by the code described in the paper. The code is available from the author upon request.

## ACKNOWLEDGEMENTS

The author thanks Lee Altenberg, Tom Blamey, Marguerite Butler, István Csabai, and Deveraux Talagi for useful comments.

## Notes

### Competing Interest Statement

The authors have declared no competing interest.

### Author Declarations

No IRB/ethics committee approval was necessary for this theoretical work.

